# The Clinical Profile of Surgically Treated Pelvic Organ Prolapse Patients in Algiers: A Retrospective Cohort Study

**DOI:** 10.64898/2025.12.13.25342186

**Authors:** Mohamed Boulahia

## Abstract

**Background:** Pelvic organ prolapse (POP) is a common condition among multiparous and postmenopausal women. In North Africa, delayed gynecological consultation often results in presentation at advanced stages requiring surgical management.

**Objective:** To describe the clinical profile, surgical management, and short-term postoperative outcomes of women undergoing surgery for pelvic organ prolapse at a tertiary care hospital in Algiers, Algeria.

**Methods:** A retrospective descriptive cohort study was conducted including 31 women who underwent surgical treatment for POP between January 2022 and December 2023. Data collected from medical records included age, parity, menopausal status, type and stage of prolapse, presenting symptoms, surgical procedures performed, and early postoperative outcomes. Descriptive statistics were applied.

**Results:** The mean age was 63.2 years (range 47–80), and the mean parity was 5.7. Most patients were postmenopausal (87%). Multicompartment prolapse was observed in 87% of cases, with stage III prolapse predominating. Vaginal hysterectomy with anterior and posterior colporrhaphy (VH + APR) was the most frequently performed procedure (58%). Early postoperative outcomes were favorable, with 90% of patients experiencing no major complications. Minor complications included transient urinary retention and mild infections, managed conservatively. The average hospital stay ranged from 3 to 5 days.

**Conclusion:** In this cohort, pelvic organ prolapse predominantly affected older, multiparous, postmenopausal women and commonly presented at advanced stages. Vaginal surgical management, particularly VH + APR, was associated with satisfactory short-term outcomes. These findings highlight the importance of preventive strategies and earlier gynecological consultation in resource-limited settings.

## Introduction

Pelvic organ prolapse (POP) is a common gynecological condition characterized by the descent of pelvic organs due to weakening of the pelvic floor support structures. It predominantly affects multiparous and postmenopausal women and can significantly impair quality of life through symptoms such as pelvic pressure, urinary dysfunction, and sexual discomfort [1,4,17].

The reported prevalence of POP varies widely depending on diagnostic criteria and study populations, with estimates ranging from mild anatomical prolapse detected on examination to symptomatic disease requiring medical or surgical intervention [1,2,10,12].Established risk factors include high parity, advancing age, vaginal childbirth, menopause, obesity, and conditions associated with increased intra-abdominal pressure [3,14,15].

In low- and middle-income countries, including North African settings, sociocultural barriers, limited access to specialized care, and reduced awareness of pelvic floor disorders may contribute to delayed presentation and advanced-stage disease at the time of diagnosis [5,6,7]. Consequently, surgical management remains the primary treatment option for many women presenting to tertiary care facilities.

Despite the clinical burden of POP, data describing the characteristics and management of surgically treated cases in Algeria remain scarce. Descriptive studies from regional hospital settings are important to document disease patterns, surgical practices, and short-term outcomes, and to inform future preventive and healthcare planning strategies.

The aim of this study was to describe the clinical profile, surgical management, and early postoperative outcomes of women undergoing surgery for pelvic organ prolapse at a tertiary care hospital in Algiers, Algeria.

## Materials and Methods

### Study Design and Setting

This was a retrospective descriptive cohort study conducted in the Department of Obstetrics and Gynecology at a tertiary care hospital in Algiers, Algeria. The study included women who underwent surgical management for pelvic organ prolapse between January 2022 and December 2023.

### Study Population

A total of 31 women who underwent surgical treatment for pelvic organ prolapse during the study period were included.

#### Inclusion criteria were

- Diagnosis of pelvic organ prolapse established by a gynecology specialist
- Surgical management performed during the study period
- Availability of complete medical and operative records

#### Exclusion criteria were

- Incomplete or missing medical records
- Non-surgical management of prolapse
- Associated gynecological malignancy

### Clinical Assessment

Pelvic organ prolapse was diagnosed clinically in the lithotomy position using the Valsalva maneuver. Staging was assigned according to the Pelvic Organ Prolapse Quantification (POP-Q) system [2,9,18]. While all patients were assigned an overall Stage (I–IV), specific point measurements (e.g., Ba, Bp, C, D) were not recorded for all subjects due to the retrospective nature of the study; therefore, data are presented as overall stages. Patient symptoms, including quality of life and sexual function, were assessed through standardized clinical interviewing during the preoperative consultation, though validated scoring instruments (such as PFDI-20) were not utilized [17].

### Data Collection

Data were extracted retrospectively from medical records and operative reports. Variables collected included age, parity, menopausal status, occupation, presenting symptoms, type and stage of prolapse, surgical procedures performed, and early postoperative outcomes.

Postoperative complications and length of hospital stay were also recorded.

### Surgical Management

All patients underwent vaginal surgical management [22]. The choice of surgical procedure was based on the type and severity of prolapse, patient characteristics, and surgeon preference. The most frequently performed procedure was vaginal hysterectomy with anterior and posterior colporrhaphy (VH + APR) [1,24]. Additional procedures included anterior or posterior colporrhaphy alone and perineorrhaphy or vault repair when indicated.

### Outcome Measures

The primary outcomes were the types of surgical procedures performed and early postoperative outcomes, including perioperative complications and length of hospital stay. Only short-term outcomes during the initial postoperative period were assessed.

### Statistical Analysis

Data analysis was performed using descriptive statistics. Continuous variables were summarized as means and ranges, while categorical variables were expressed as frequencies and percentages.

### Surgical Technique

All patients underwent vaginal surgical management under regional or general anesthesia. Surgical procedures were selected based on the type and severity of pelvic organ prolapse, intraoperative findings, patient characteristics, and surgeon preference.

### Vaginal Hysterectomy and Apical Support

A standard vaginal approach was utilized. Following circumferential incision and bladder dissection, the pedicles were secured using the Heaney technique. To ensure apical support and prevent future vault prolapse, a modified McCall culdoplasty was performed in all cases undergoing hysterectomy, incorporating the uterosacral ligaments into the vaginal vault closure [24].

### Decision Criteria for Additional Procedures

The decision to perform perineorrhaphy was based on a measured perineal body of <2 cm or the presence of significant levator ani diastasis. Anti-incontinence procedures (e.g., Kelly plication) were performed only in patients with masked stress urinary incontinence (SUI) demonstrated intraoperatively after prolapse reduction [21,22]. Systematic mid-urethral slings were not performed due to the lack of preoperative urodynamic confirmation.

### Anterior Colporrhaphy

Anterior vaginal wall repair was performed for cystocele correction. Following a midline incision of the anterior vaginal mucosa, the vaginal epithelium was dissected from the underlying bladder. The pubocervical fascia was plicated in the midline using delayed absorbable sutures to restore anterior compartment support, followed by trimming and closure of the vaginal mucosa.

### Posterior Colporrhaphy

Posterior repair was performed in cases of rectocele. A midline posterior vaginal incision was made, and the vaginal mucosa was separated from the rectovaginal fascia. The rectovaginal fascia was reinforced by midline plication, and excess vaginal mucosa was excised before closure [13].

### Anti-incontinence Procedures

Concomitant anti-incontinence procedures were not routinely performed. Preoperative urodynamic testing and validated stress urinary incontinence assessments were not systematically available during the study period, and surgical management focused on correction of symptomatic prolapse. Patients with postoperative urinary symptoms were managed conservatively and followed clinically.

### Ethical Considerations

This study was approved by the institutional ethics committee of Hassen Badi Hospital El Harrach. Due to the retrospective design and use of anonymized data, informed consent was waived, in accordance with the Declaration of Helsinki.

## Results

### Epidemiological Characteristics

A total of 31 women underwent surgical management for pelvic organ prolapse during the study period. The mean age was 63.2 years (range 47–80 years), with the highest proportion of patients aged between 60 and 69 years. Most patients were postmenopausal (87%). Multiparity was common, with a mean parity of 5.7. The age distribution of patients is summarized in **Table 1**.

**Table 1.** Demographic and Epidemiological Profile (n=31)

| Characteristic | Value (Mean ± SD or n/%) |
| --- | --- |
| Age (Years) | 63.2 ± 8.4 |
| Mean Parity | 5.7 ± 1.2 |
| Postmenopausal Status | 27 (87.1%) |
| Body Mass Index (BMI) | 28.4 ± 4.1 kg/m<sup>2</sup> |
| History of Home Delivery | 22 (71.0%) |

### Clinical Characteristics

Multicompartment prolapse was the most frequent presentation, observed in 87% of patients. The majority of cases were classified as stage III prolapse at the time of surgery. Common presenting symptoms included pelvic heaviness or vaginal bulge sensation (71%), urinary symptoms (52%), and sexual discomfort (35%). Clinical characteristics are summarized in **Table 2**.

**Table 2.** Clinical characteristics and surgical management of pelvic organ prolapse (n = 31)

| Variable | Frequency (n) | Percentage (%) |
| --- | --- | --- |
| **Clinical Findings** |  |  |
| – Stage III prolapse (predominant) | 28 | 90.3 |
| – Multicompartment involvement | 27 | 87.1 |
| – Sexual discomfort (symptom) | 11 | 35.5 |
| **Surgical Procedures** |  |  |
| – VH + APR + McCall culdoplasty | 18 | 58.1 |
| – Isolated anterior colporrhaphy | 6 | 19.4 |

### Surgical Procedures

All patients underwent vaginal surgical management. Vaginal hysterectomy with anterior and posterior colporrhaphy (VH + APR) was the most commonly performed procedure (58%). Other surgical interventions included anterior colporrhaphy alone (19%), posterior colporrhaphy alone (13%), and perineorrhaphy or vault repair (10%). Surgical procedures are summarized in **Table 2**.

### Postoperative Outcomes

Early postoperative outcomes were favorable in most patients. Twenty-eight women (90%) experienced no major complications. Minor complications included transient urinary retention and mild postoperative infections, which were managed conservatively. No major hemorrhage, visceral injury, or early recurrence was recorded during the initial postoperative period. The average length of hospital stay ranged from 3 to 5 days.

### Visual Summary of Findings

Figure 1 provides a visual overview of patient characteristics, disease severity, and outcomes. It emphasizes the prevalence of advanced-stage disease and the clinical efficacy of the vaginal surgical approach in this population.

**Figure 1.**
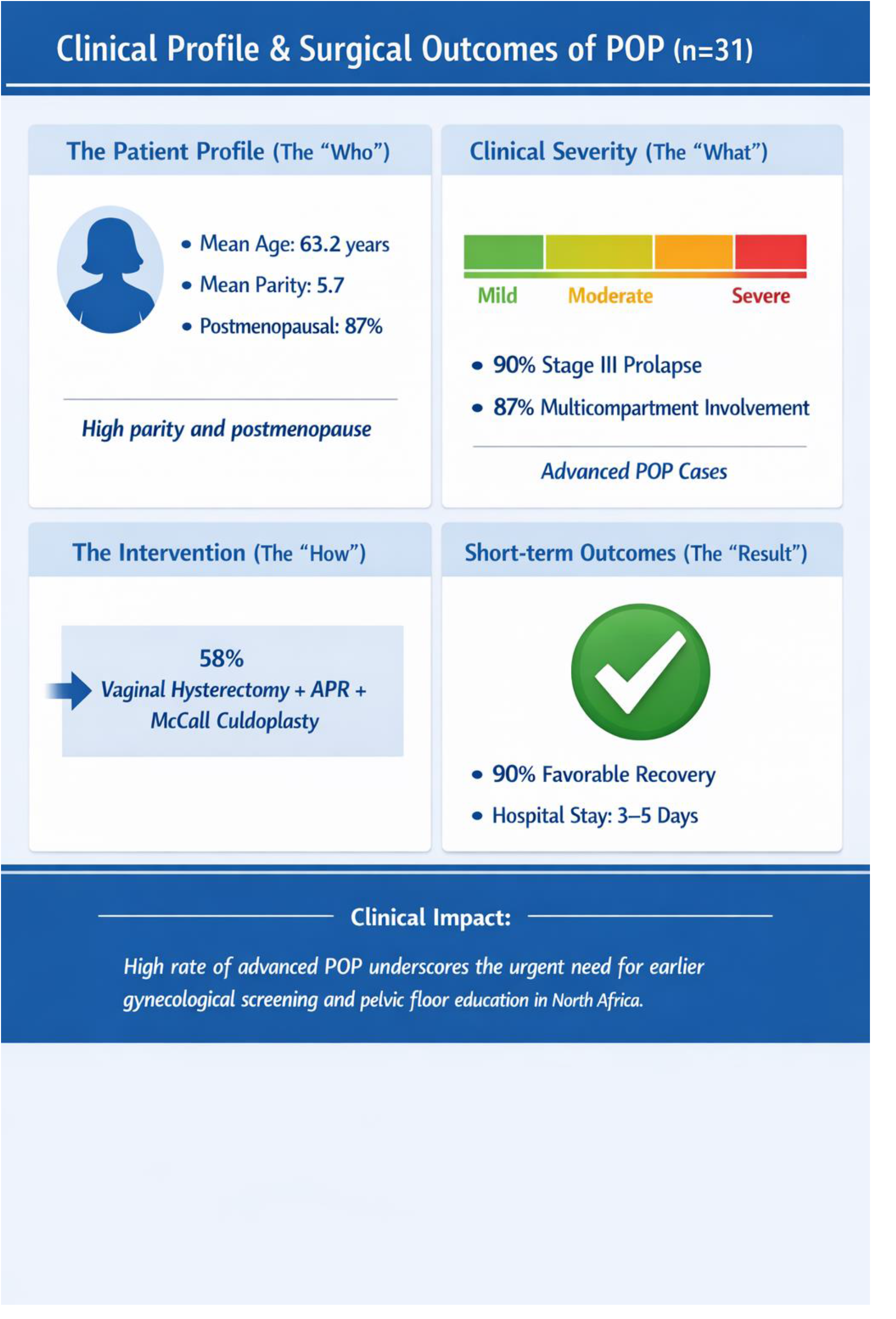
Clinical profile and surgical management of POP in Algiers (n=31). Infographic summarizing the predominance of Stage III multicompartment disease in multiparous women and the favorable outcomes associated with vaginal surgery and McCall culdoplasty.

## Discussion

### Main Findings

This retrospective cohort study describes the clinical profile and surgical management of women treated for pelvic organ prolapse at a tertiary care hospital in Algiers, Algeria. The findings indicate that POP predominantly affected older, multiparous, postmenopausal women and was most commonly diagnosed at an advanced stage. Multicompartment prolapse was the predominant presentation, reflecting delayed healthcare-seeking behavior and limited access to early gynecological assessment.

These findings are consistent with reports from other North African and South Asian settings, where high parity, menopause, and advancing age are established risk factors for POP and late presentation is common [5,6,7]. In contrast, studies from high-income countries often report earlier-stage diagnosis, likely reflecting greater awareness, preventive care, and routine gynecological follow-up [8,10,12].

### Surgical Management and Outcomes

Vaginal surgery was the exclusive approach used in this cohort, with vaginal hysterectomy combined with anterior and posterior colporrhaphy (VH + APR) being the most frequently performed procedure. This approach remains widely used in resource-limited settings due to its effectiveness, familiarity, and avoidance of specialized equipment [1,22]. In the present study, short-term postoperative outcomes were favorable, with a low rate of minor complications and no major intraoperative events or early recurrences.

Although long-term outcomes and objective functional measures were not assessed, the observed early postoperative course aligns with existing literature supporting vaginal repair as a safe and effective option for advanced prolapse when uterine preservation is not a priority [9,24].

### Strengths and Limitations

A key strength of this study is its focus on a population for which published data on surgically treated POP are limited. By reporting real-world surgical practice and early outcomes in a tertiary hospital setting, this study contributes baseline information relevant to similar healthcare contexts.

Several limitations should be acknowledged. The study was retrospective, single-center, and included a relatively small number of patients, which limits generalizability. Detailed POP-Q measurements and validated symptom or quality-of-life questionnaires were not consistently available, precluding quantitative anatomical and functional outcome assessment. Additionally, only short-term postoperative outcomes were evaluated, and long-term recurrence and functional results could not be assessed, the lack of preoperative urodynamic testing and long-term follow-up are noted limitations [19,20].

While our cohort size is limited (n=31), it represents a specific surgical window in an Algiers tertiary center. The high prevalence of Stage III disease and multicompartment involvement (87%) highlights a significant delay in seeking care compared to Western cohorts. This study serves as a necessary baseline for pelvic floor health in the region, where sociocultural factors often lead to patients presenting only when surgical intervention is the only remaining option.

### Clinical and Public Health Implications

Despite these limitations, the findings highlight important clinical and public health considerations. The predominance of advanced-stage prolapse underscores the need for improved awareness, early detection, and preventive strategies. Pelvic floor muscle training, optimized obstetric care, family planning education, and timely gynecological consultation may reduce the incidence of severe prolapse and the need for complex surgical intervention.

From a surgical perspective, the continued reliance on vaginal procedures reflects their practicality and effectiveness in resource-limited settings. Standardized preoperative assessment and incorporation of validated outcome measures could further enhance care quality and future research.

### Future Research

Future studies should prioritize the establishment of larger, multicenter cohorts that incorporate standardized POP-Q measurements and validated quality-of-life instruments (such as the PFDI-20). Long-term follow-up is essential to evaluate the durability of vaginal repairs and to accurately document recurrence rates in the North African population. Furthermore, community-based prevalence studies are needed to guide national health policy and optimize resource allocation for pelvic floor disorders in Algeria.

Beyond clinical outcomes, research should evolve to explore the underlying genetic risk factors specific to our region. Identifying these hereditary predispositions could facilitate the transition toward personalized screening and preventive strategies, allowing for earlier intervention in women at high genetic risk for advanced-stage prolapse [25].

## Conclusion

This retrospective cohort study provides a descriptive overview of women undergoing surgical management for pelvic organ prolapse at a tertiary care hospital in Algiers. Pelvic organ prolapse predominantly affected older, multiparous, postmenopausal women and commonly presented at an advanced stage, reflecting delayed consultation and limited access to early gynecological care. Multicompartment involvement was frequent, and vaginal surgical approaches were the mainstay of treatment.

Vaginal hysterectomy with anterior and posterior colporrhaphy was the most commonly performed procedure and was associated with favorable short-term postoperative outcomes and a low rate of minor complications. Although long-term outcomes and validated functional measures were not assessed, these findings support the continued role of vaginal surgery as a practical and effective option for advanced prolapse in resource-limited settings [1, 24]. The high prevalence of advanced-stage disease highlights the need for preventive strategies, including pelvic floor muscle training, improved obstetric care, family planning education, and earlier gynecological consultation. Future multicenter studies incorporating standardized anatomical assessment and long-term follow-up are needed to better inform clinical practice and health policy in similar settings [11, 15, 25].

## Data Availability

The datasets generated and/or analyzed during the current study are no longer available, as the retrospective data collection was anonymized and not retained beyond the study period. All results derived from these data are fully reported within the manuscript.

## Acknowledgments

I would like to acknowledge the support of colleagues at Hassen Badi Hospital El Harrach for access to data.

## Author Contributions (CRedIT Statement)

Mohamed Boulahia: conception, data collection, analysis, drafting, and approval of final manuscript.

## Ethical Approval

This study was approved by the institutional ethics committee of Hassen Badi Hospital El Harrach. Formal approval was waived due to the retrospective nature of anonymized data collection, in accordance with the Declaration of Helsinki.

## Consent to Participate

Patient consent was waived due to retrospective anonymized data.

## Presentation/Awards

None.

